# Factors associated with risk of developmental delay in preschool children in a setting with high rates of malnutrition: a cross-sectional analysis of data from the IHOPE study, Madagascar

**DOI:** 10.1101/19011064

**Authors:** Ann C. Miller, Andres Garchitorena, Faramalala Rabemananjara, Laura Cordier, Marius Randriamanambintsoa, Victor Rabeza, Hery-Tiana Rahaniraka Razanadrakoto, Ranaivozafindary Rakoto Ramakasoa, Olivier RamahefarisonTiana, Baolova Nathaline Ratsimbazafy, Mohammed Ali Ouenzar, Matthew H. Bonds, Lisy Ratsifandrihamanana

## Abstract

Nearly two hundred eighty million children in low and middle income countries are at risk for not achieving their full developmental potential(1). Poor development can lead to reduced scholastic ability(2) or opportunity (promoting and entrenching inequity), and has been associated with lower adult earning potential(3), and poorer health(4), thus potentially contributing to intergenerational poverty traps.

While the entire period of childhood is important for development, the first 1000 days (conception to age 2) are critically important for brain development; with the most rapid and prolific development of neural pathways, first in sensory development, then language skills and then higher cognitive functioning(5). Experiences during this period – positive or negative – can substantially impact the architecture of the developing brain. Adverse events such as exposure to familial or societal violence, toxins such as lead or arsenic, illness (anemia, diarrhea, HIV, chronic malnutrition, maternal depression), or lack of a developmentally stimulating and nurturing environment can all act as toxic stressors on young children that negatively impact cognitive development throughout the life-course(6). Infants and young children in settings of poverty are often faced with many of these stressors(7).

---

Some of these factors, if identified early enough, are potentially modifiable through intervention, such as caregiver interaction and stimulation(8, 9), or treatment for or prevention of malnutrition(3, 10). System and policy changes can influence access to health care for prevention and treatment of illness, malnutrition, early marriage and first birth, and maternal education, all of which may influence children’s development. However, an assessment of context-specific risks is necessary to develop interventions that improve child development in specific settings.

Madagascar is one of the poorest nations in the world, with 75.1% of the population living on less than $1.90 USD in 2018. It has very high rates of developmental risk (both as measured through direct assessment in household surveys and indirectly through rates of stunting) and of many possibly modifiable risk factors. In surveys from 2012 (most recent estimates to date), an estimated 32% of children ages 3-5 years were considered developmentally at risk, 8.5% received developmental stimulation from an adult and 47% of children under age 5 were stunted (11, 12); the fifth highest prevalence of stunting in the world. In early 2017, to address the burden of stunting and attendant neurodevelopmental delay, Madagascar unveiled their third National Nutrition Action Plan (PNAN III), a multisector proposal co-funded by the World Bank that aims to reduce stunting in Madagascar from 47.3% to 38% by 2021. The PNAN III plan includes system improvements to deliver nutritional and health services in 8 regions of high stunting; with support for other sectors, such as water, sanitation and hygiene, and climate-smart agriculture (13). However, minimal information exists from these regions about the baseline burden of risk or determinants for developmental delay in children, which could help inform the implementation of PNAM-III and other health system strengthening interventions.

In 2014, the nongovernmental health organization, PIVOT, began operations in partnership with the Ministry of Health to establish Ifanadiana District, in Southeastern Madagascar, as a model health system. The effort aimed to strengthen the public health system through improvements in facility readiness, clinical programs, and integrated data systems at all levels of care. That year, we initiated the Ifanadiana Health Outcomes and Prosperity longitudinal Evaluation (IHOPE), a representative cohort of Ifanadiana District to baseline health and socio-economic status of the population and monitor the impact of the health intervention on the population over time. Here, we use data from the 2016 wave of the IHOPE cohort at the time of the initiation of PNAN-III to quantify the burden of risk for developmental delay in children ages 3 to 5 years in Ifanadiana District, and to determine potentially modifiable risk factors that could inform local program development.

## Methods

### Study population

Our study population is based in Ifanadiana District, with a population of approximately 209,000 residents. Ifanadiana is a low-resource setting in a low-resource country. Approximately 85% of the population subsist from agriculture, 3% have access to improved latrines and only 20% have access to safe drinking water. Health indicators are low compared with Madagascar as a whole—in 2014, 34% of children had all appropriate vaccines by the time they were 23 months old (vs. 51% for Madagascar); under 5 mortality was estimated at 145/1000 live births (vs. 62/1000 for Madagascar); and 20% of mothers had trained assistance in delivering their last baby (vs. 59% for Madagascar). Children’s development in the district is similarly low compared to the country as a whole. Four percent of children ages 3 or 4 were attending preschool (vs. 7.7% nationally) and 27% of adults between ages 18-35 had no formal education (vs. 21% nationally)

### Data collection

The IHOPE cohort was established as an extension of a representative baseline survey from 2014, which occurred at the outset of the health system strengthening intervention. The survey (14) used a 2-stage random sample in 80 clusters to select 1600 households. The IHOPE study revisits the same households every second year to collect data on health and economic indicators, and it is stratified to be representative of both within and outside the initial PIVOT catchment area (15). The IHOPE study uses methods, techniques and questionnaires based primarily on Demographic and Health Surveys (DHS), with additional questions from the Multiple Indicator Cluster Surveys (MICS), and the Rwanda Questionnaire on Well-being (16). Specifically, we used the early child development index (ECDI) modules from the MICS 4. The ECDI is designed to be an internationally comparable population screener for developmental risk(17, 18) (as opposed to a diagnostic tool for developmental delay), which includes questions about development of 3 and 4 year olds, including risk and protective factors. Specific questions include whether adults interact with the child in developmentally supportive ways (reading, counting or naming things, singing to the child, playing with the child, taking the child out of the home compound) (See Table 1 for specific questions in the ECDI). The ECDI focuses on 4 domains—learning, physical growth, social-emotional and literacy/numeracy. The IHOPE surveys are implemented and conducted by the Madagascar Institute of Statistics (INSTAT), the same organization that conducts the DHS and MICS in Madagascar.

**Table 1:** ECDI domains, questions and criteria for “on-track” development

| Domain | Questions | Criteria for "on track" development |
| --- | --- | --- |
| Literacy/numeracy | Can [Name] identify/name at least 10 letters of the alphabet? | On track if at least 2 are positive. |
|  | Can [Name] read at least 4 simple, popular words? |  |
|  | Does [Name] know the name and recognize the symbol of all numbers from 1 to 10? |  |
| Physical | Can [Name] pick up a small object with 2 fingers, like a stick or a rock from the ground? | On track if at least 1 is positive |
|  | Is [Name] sometimes too sick to play? (*on track if false*) |  |
| Social Emotional | Does [Name] get along well with other children? | On track if at least 2 are positive. |
|  | Does [Name] hit, kick or bite other children or adults? (*on track if false*) |  |
|  | Does [Name] get distracted easily? (*on track if false*) |  |
| Approaches to Learning | Does [Name] follow simple directions on how to do something correctly? | On track if at least 1 is positive |
|  | When given something to do, is [Name] able to do it independently? |  |
| Total ECDI |  | Percentage of children on track in 3/4 domains. |

### Definitions

#### Outcomes

We defined development outcomes in 2 ways; one to compare to external cohorts in other countries, and one to consider risks to development in children within the cohort relative to one another. For external comparisons, we used the standard definitions from the MICS. Using these definitions, a child was considered to be developmentally “on track” if the child had a positive score in three of four domains in the ECDI, and “at risk for delay” if positive scores in fewer than 3 domains. For internal comparisons, a child was considered to have “low development relative to peers” if the ECDI score was at least 1 standard deviation (SD) below the IHOPE sample mean.

#### Stimulation indicators

We used the standard definitions in the MICS to create our stimulation indicators. A variable for “adult involvement” was created as a continuous score of number of reported developmentally stimulating activities conducted with a child by any adult (mother, father or other adult). We also created a binary variable for involvement—”adult disengagement” if no adult did at least 4 learning activities with the child in the last 3 days, and maternal, paternal and “other adult” disengagement to assess differences between caregivers. Home environment was assessed with a series of specific questions about number and source of playthings, number of books in the house, and whether and how long the child was left alone or with another child under the age of 10 years as a caregiver (Figure 1).

#### Other Child-level measures

Nutritional status was assessed using the World Health Organization measures(19); both continuous weight-for-age (underweight), weight-for-height (wasting) and height-for-age (stunting) age-adjusted z scores compared to a normed international population, and binary variables to characterize moderate and severe stunting, wasting and underweight status.

#### Other household-level measures

The number of other children under age 5 in the house was assessed as a continuous variable. A binary variable was created for maternal education—any years of formal education vs. no formal education. Household poverty was assessed using wealth indices as determined using DHS methods using principal components analysis(20). Cutoff points for wealth quintiles were determined to be the values closest to but less than the 20, 40, 60 and 80^th^ percentiles of the cumulative wealth index. We estimated for all household members, injury or illness in the 4 weeks prior to the interview, whether household members had sought care for these illnesses or injuries, and whether they had had to miss work activities or school because of the illness/injury. Maternal age at birth was calculated from estimated maternal and child birthdates and was assessed as a continuous variable and a 4-category variable (age 15-19, 20-24, 25-34 and 35-49) Orphan status was presented as a categorical variable (single maternal; single paternal; both; neither; and unknown status if one or other parent’s survival was unknown); and fathers’ presence in the home was assessed as a binary variable.

### Data analysis

We conducted descriptive analysis using frequencies and percentages for binary and categorical data and medians with interquartile ranges for continuous data. We analyzed data using Stata 15 (Stata: College Station, TX). We assessed factors associated with the outcomes (risk for delay or low development related to peers) usinglogistic regressions (for binary endpoints) and accounting for clustering at the sampling level using Stata’s “cluster” function. Factors in univariable analysis significant at an alpha of 0.1 were entered into a full multivariable model and reduced to a final model using backward stepwise regression with an alpha of 0.05 for retention. Wald tests were used to compare reduced to full models. We assessed factors in the final models for interaction.

### Ethics review

The protocol and tools for the IHOPE cohort were reviewed and approved by the Harvard Medical School Institutional Review Board and the Madagascar National Ethics Committee. Verbal consent was obtained from eligible adults (ages 15-59) and from parents or legal guardians for children’s participation. All data was de-identified data prior to analysis; investigators had only access to data identifiable at the cluster level.

## Results

### Sample description

432 children from the 2016 wave met eligibility criteria for inclusion (ages 3 to <5 years, alive at interview). Table 2 describes demographics, anthropometry and developmental status of the study population. Of the 432 children included, 259 (59.9%) were considered to be developmentally on track, and 173 (40.1%) were at risk for delay based on ECDI scores. Sixty-eight children (16.0%) were considered to have low development compared to peers. In our sample, the mean ECDI score was 5.12 (range 1-10, SD 1.56). The mean for children at risk for delay was 4.6 (1.8 SD) and 5.5 (1.23 SD) in children whose development was on track for both outcome measures. For children with low development compared to peers, mean ECDI score was 2.8(0.57 SD). By individual domain, literacy/numeracy had the lowest proportion (7.2%) of children with on-track development, while physical (85.4%), learning (86.6%) and socio-emotional (82.6%) domains had substantially higher proportions.

**Table 2:** Descriptive statistics of the population of children ages 3-5 years old in the 2016 wave of the IHOPE study, Madagascar

|  |  |  |
| --- | --- | --- |
| **Demographics** |  |  |
| Male sex | 212 | 49 |
| Age |  |  |
| 3 | 202 | 47.2 |
| 4 | 228 | 52.8 |
| Number of children under 5 in household |  |  |
| 1 | 157 | 36.3 |
| 2 | 215 | 49.8 |
| 3 | 50 | 11.6 |
| 4 | 8 | 1.8 |
| 5 | 2 | 0.5 |
| Wealth quintile |  |  |
| poorest | 96 | 22.2 |
| second-poorest | 109 | 25.2 |
| middle | 105 | 24.3 |
| second-richest | 67 | 15.5 |
| richest | 55 | 12.7 |
| Mean (SD) anthropometrics<br>(n=400) |  |  |
| Height for age z | -2.04 | 1.3 |
| Weight for age z | -1.66 | 0.91 |
| Weight for height | -0.57 | 0.94 |
| Mother's age |  |  |
| 15-19 | 23 | 5.3 |
| 20-24 | 116 | 26.8 |
| 25-34 | 169 | 39.1 |
| 35-49 | 124 | 28.7 |
| Orphan status |  |  |
| Single, mother deceased | 1 | 0.23 |
| Single, father deceased | 15 | 3.6 |
| Single, father status unknown | 10 | 2.3 |
| Double | 0 |  |
| Not orphan | 406 | 93.87 |
| Maternal education |  |  |
| No formal | 162 | 37.5 |
| Primary | 238 | 55.1 |
| Secondary or higher | 32 | 7.4 |
| **Development indicators** |  |  |
| At-risk (ECDI scores in <3/4) | 173 | 40.1 |
| Mean ECDI score | 5.13 | 1.54 |
| On track in physical domain | 369 | 85.4 |
| on track in learning | 374 | 86.5 |
| On track in social-emotional domain | 357 | 82.6 |
| On track in literacy/numeracy domain | 31 | 7.2 |
| Low development compared to peers ($Z < 0.1$) | 68 | 15.8 |
| Mean ECDI score in low development | 2.76 | 0.55 |
| Adult involvement |  |  |
| The child was engaged in 4 or more stimulation activities with adults in last 3 days | 76 | 17.6 |
| Mother engaged in 4 or more activities n=431 | 9 | 2.1 |
| Father engaged I 4 or more activities n=431) | 1 | 0.23 |
| Other, non-parental adult engaged in 4 or more activities n=431 | 26 | 6.03 |
| Median number of activities | 2 | range 0-8 |
| Median number for mothers | 1 | range 0-5 |
| Mean number for fathers | 0 | range 0-4 |
| Mean number for other adults | 0 | range 0-6 |
| Number of children with no supporting activities | 106 | 24.6 |

Of 431 children with at least some completed developmental support questions,76 (17.6%) had adult support for development (4 or more different developmentally stimulating activities in the last 3 days), and 106 children (24.6%) had no adult support for development (0 developmentally supportive activities conducted by an adult in the last 3 days). The median number of developmentally supportive activities provided to a child was 2 [range 0-8].

Factors in univariable analysis associated at an alpha of 0.1 with being at risk for delay compared to international standards included household poverty (bottom 2 quintiles—Odds Ratio (OR) 1.52 95% Confidence Interval CI (1.03, 2.25)), adult disengagement, and increasing number of activities provided by the father (OR 1.39 95%CI 1.04, 1.86). Factors associated with being developmentally on track included being left in the care of an older child (OR for delay 0.48 95%CI 0.30,0.76), an adult playing (OR 0.49 95%CI 0.31, 0.76) or singing songs (OR 0.58 95%CI0.37. 0.89) with the child, and higher number of activities by a non-parental adult (OR 0.64 95% CI 0.53, 0.76). Table 3 shows full and reduced models of factors associated with being at risk for delay. In the final model, increase in number of activities by the father (adjusted OR (aOR)1.59 95% CI1.13, 2.21) was associated with being at risk, and adult playing with the child (aOR 0.54 95%CI 0.31, 0.94) and increase in activities provided by a non-parental adult (aOR 0.64 95% CI 0.53, 0.76) were associated with on track development.

**Table 3:**
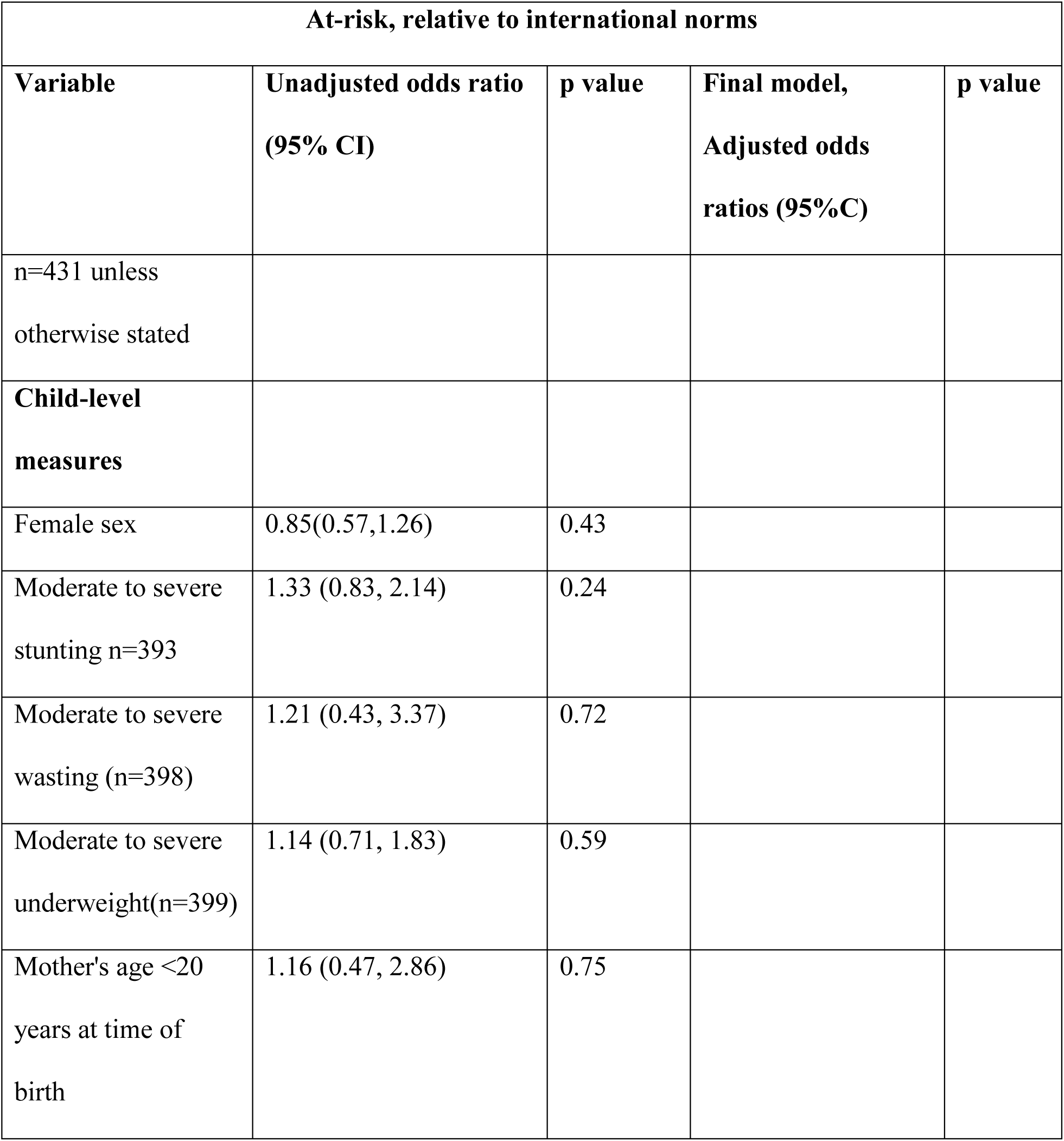
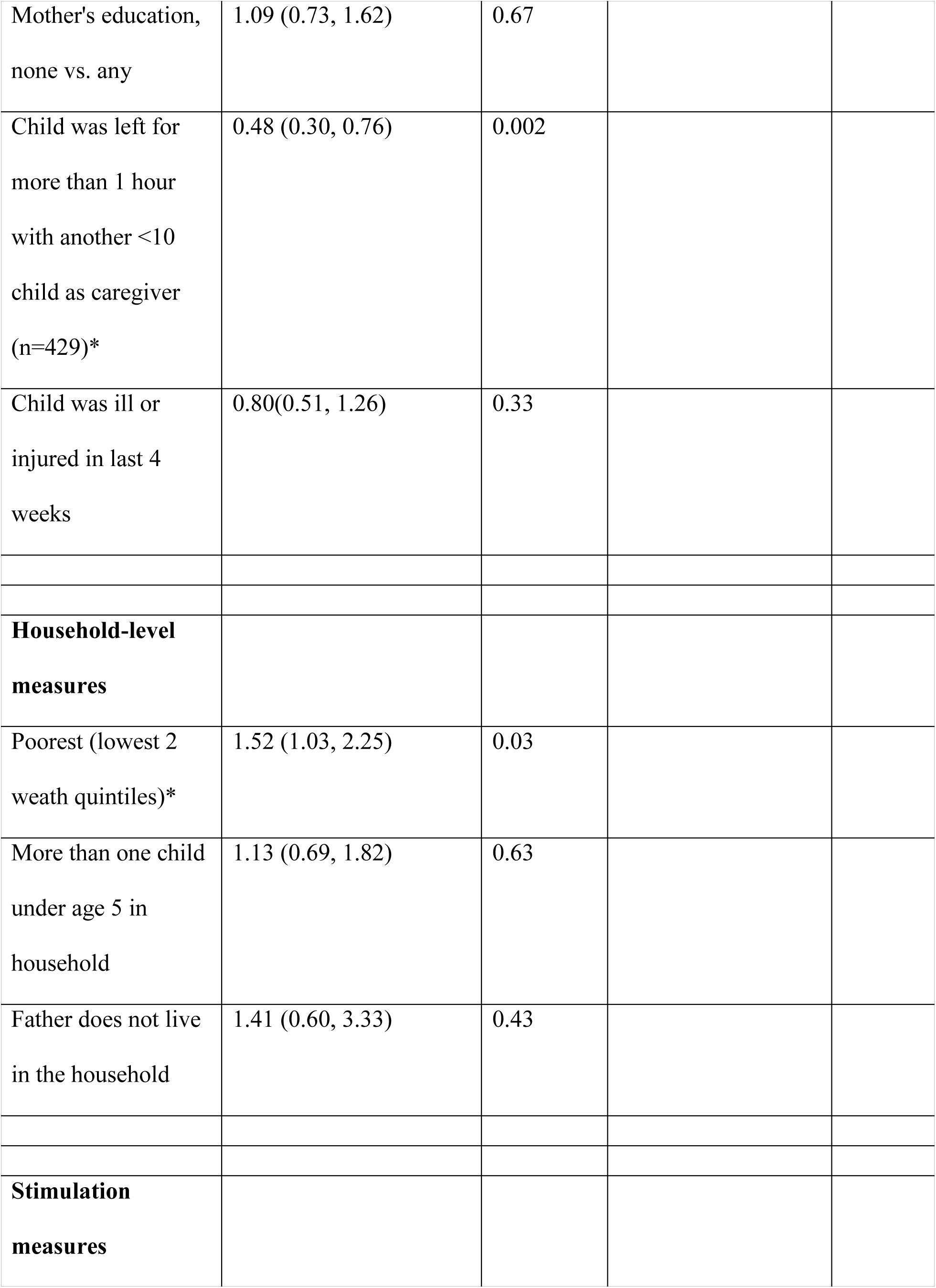
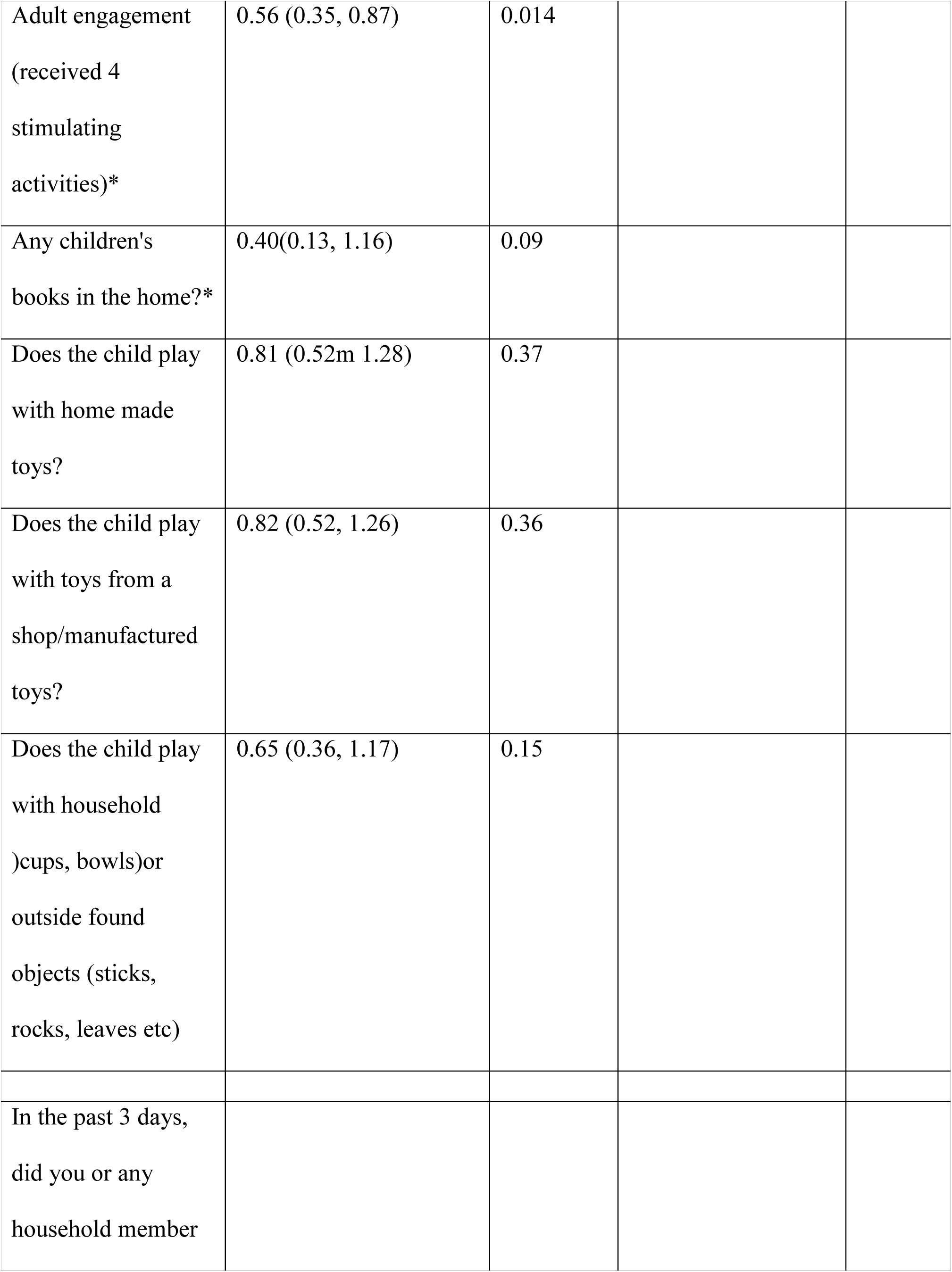
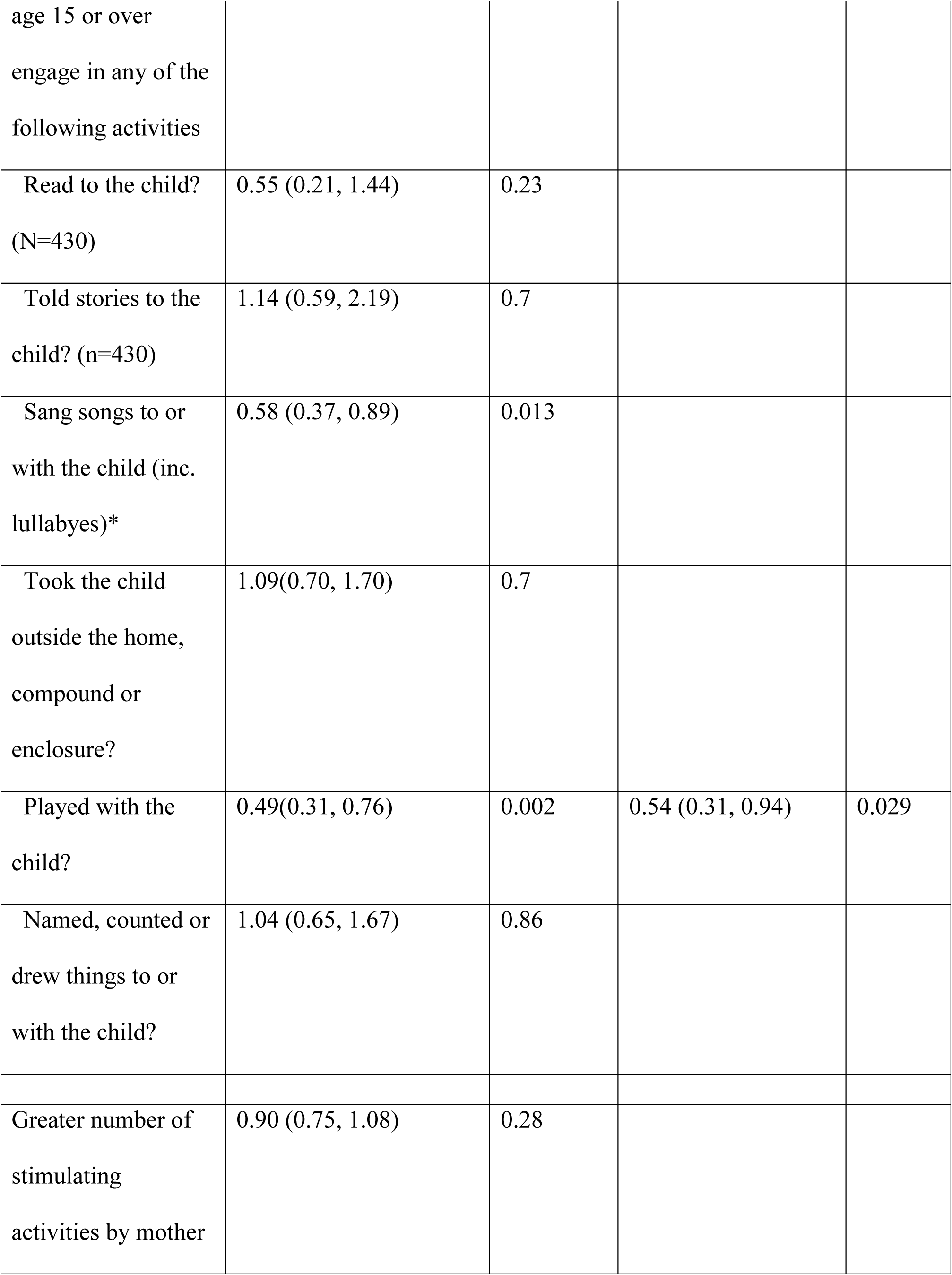
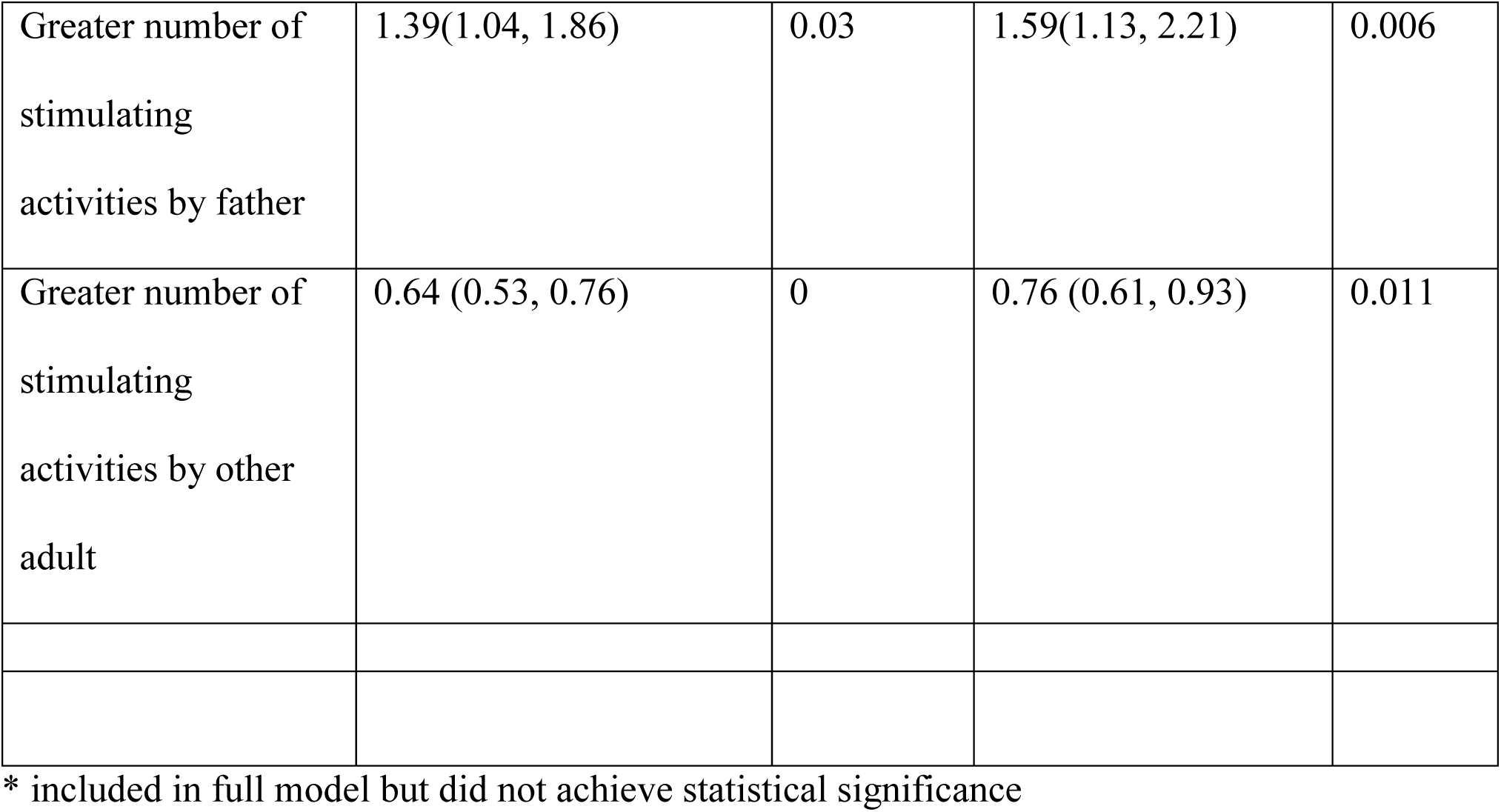
Univariable and multivariable models of factors associated with developmental risk in children ages 3-5 years in rural Madagascar, 2016. Comparison with international reference population.

Table 4 shows factors in the full and reduced models for low development compared to peers. In univariable analyses, factors negatively associated at an alpha of 0.1 with development compared to peers included household poverty (bottom 2 wealth quintiles OR 2.16 95%CI 1.20, 3.89)), adolescent mother (age <20) (OR 3.09 95%CI1.18, 8.13), an increasing number of other under 5 children in the home (OR 2.06 95%CI1.11, 3.82), and having moderate to severe wasting (OR 2.65 95%CI 0.91, 7.72)). Factors positively associated with development included having an adult engage in at least 4 developmentally supportive activities (OR 0.33 for delay 95%CI 0.15, 0.72), increase in engagement by a non-parental adult (OR 0.43 95%CI0.30, 0.63), playing with household or other found objects as toys (OR 0.41 95%CI 0.21, 0.79), specific activities of an adult playing with (OR 0.49 95%CI 0.25, 0.97) and singing to the child (OR 0.44 95%CI 0.24, 0.81), the child having an illness or injury in the last 4 weeks (OR 0.43 95%CI 0.24, 0.81) or being left with another child as caregiver (OR 0.45 95%CI0.21, 0.94). In the final model, having other under 5 children in the household, an adolescent mother (aOR 3.89 95%CI1.32, 11.48), and an adult taking the child outside (aOR 3.03 95%CI 1.14, 6.45) were associated with having low development scores. Being ill or injured in the last 4 weeks (aOR for delay 0.40 95%CI 0.21, 0.76), playing with household or found objects as toys (aOR 0.33 95%CI 0.14, 0.73) and an increase in activities with an adult other than mother or father (aOR 0.2895%CI0.16, 0.50) were associated with higher development scores.

**Table 4:** Univariable and multivariable models of factors associated with developmental risk in children ages 3-5 years in rural Madagascar, 2016. Within-cohort comparison.

| Development compared to Malagasy peers |  |  |  |  |
| --- | --- | --- | --- | --- |
| Variable | Unadjusted<br>odds ratio<br>(95% CI) | p<br>value | Final model,<br>Adjusted odds<br>ratios (95%C) | p value |
| n=431 unless otherwise stated |  |  |  |  |
| **Child-level measures** |  |  |  |  |
| female sex | 0.90 (0.52,<br>1.53) | 0.69 |  |  |
| moderate to severe stunting n=393 | 0.91(0.47,<br>1.74) | 0.77 |  |  |
| moderate to severe wasting<br>(n=398)* | 2.65(0.91,<br>7.72) | 0.07 |  |  |
| moderate to severe<br>underweight(n=399) | 1.21 (0.65,<br>2.25) | 0.55 |  |  |
| mother's age <20 | 3.09(1.18,<br>8.13) | 0.02 | 3.89 (1.32, 11.48) | 0.014 |
| mother's education, none vs. any | 1.28(0.77,<br>2.14)) | 0.33 |  |  |
| left for more than 1 hour with another <10 child as caregiver<br>*(n=429) | 0.45 (0.21, 0.94) | 0.035 |  |  |
| child was ill or injured in last 4 weeks | 0.43 (0.24, 0.81) | 0.005 | 0.40(0.21, 0.76) | 0.005 |
| **Household-level measures** |  |  |  |  |
| poorest (lowest 2 wealth quintiles)* | 2.16(1.20, 3.89) | 0.01 |  |  |
| more than one under 5 child in household* | 2.06 (1.11, 3.82) | 0.021 |  |  |
| father does not live in the household | 1.76 (0.67, 4.57) | 0.25 |  |  |
| **Stimulation measures** |  |  |  |  |
| adult engagement (received 4 stimulating activities)* | 0.33(0.15, 0.72) | 0.006 |  |  |
| any children's books in the home?(n=430) | 0.88(0.24, 3.28) | 0.85 |  |  |
| does the child play with home made toys? (n=430) | 0.62(0.30, 1.28) | 0.19 |  |  |
| does the child play with toys from a shop/manufactured toys?* | 0.45 (0.19, 1.07) | 0.07 |  |  |
| does the child play with household<br>(cups, bowls)or outside found<br>objects (sticks, rocks, leaves etc) | 0.41 (0.21,<br>0.79) | 0.008 | 0.33 (0.14, 0.73) | 0.007 |
| In the past 3 days, did you or any household member age 15 or over engage in any<br>of the following activities |  |  |  |  |
| read to the child? (N=430) | 0.52(0.13,<br>1.98) | 0.34 |  |  |
| told stories to the child? (n=430) | 1.10 (0.46,<br>2.85) | 0.84 |  |  |
| sang songs to or with the child (inc.<br>lullabies)* | 0.44 (0.24,<br>0.81) | 0.009 |  |  |
| took the child outside the home,<br>compound or enclosure? | 1.82 (0.96,<br>3.44) | 0.06 | 3.03 (1.14, 6.45) | 0.004 |
| played with the child?* | 0.49(0.25,<br>0.97) | 0.041 |  |  |
| named, counted or drew things to or<br>with the child? | 0.66(0.28,<br>1.58) | 0.35 |  |  |
| greater number of stimulating<br>activities by mother | 1.14 (0.93,<br>1.40) | 0.21 |  |  |
| greater number of stimulating<br>activities by father | 1.27(0.84,<br>1.94) | 0.25 |  |  |
| greater number of stimulating<br>activities by other adult | 0.43 (0.30,<br>0.63) | 0 | 0.28(0.16, 0.50) | 0 |

462 \* included in full model but did not achieve statistical significance

## Discussion

A high proportion of the children (40.1%) were considered to be at risk for delay based on international standards. This was driven mostly by the literacy and numeracy domain, with only 7% of children considered developmentally on track. This is similar to the most recent published subnational MICS for Madagascar (2012) in which 7.4% of children were on track in this domain, compared to 94.2% in physical, 80.3 in socio-emotional and 83.7% in the learning domain(11). Early literacy in western cultures is associated with better school performance, and early language skills are associated with cognitive development in both low and high resource settings(21-23). Relative poverty has been shown in both wealthy and poor settings to be associated with lower performance on language scales. A study by Fernald et al in Madagascar demonstrated that poverty relative to others was also associated with lower development, particularly in the language domain(24). Although relative poverty was not associated with lower development in our study after adjusting for other factors including playing with the child and increase in stimulation by adults, this finding aligns with a 2000 study in the US(25) that found that effect of poverty on development was completely mediated by health and home stimulation factors. Poverty’s independent influence on development merits further study.

There may be differences in cultural norms around when the foundations of literacy are laid down. Conversations with parents in Ifanadiana suggest that although physical and socio-emotional development are the focus of attention by caregivers, children are not expected to learn to read until they start public school at age 5. Parents in Ifanadiana often have low literacy themselves, with limited capacity to prioritize and model literacy and numeracy for young children outside of school. And culturally, tools to stimulate development of this domain, such as children’s picture books, are both very rare in Malagasy languages, and very expensive to obtain (personal communication, LRR). Another possible explanation of this phenomenon is that the ECDI literacy/numeracy domain is simply inappropriate in this setting. The numeracy component in particular, asks if children know the name and recognize the symbols of numbers up to 10, which is more advanced for the same age on comparable tools, which assess a child’s ability to simply count to 10. (McCoy).. The use of the ECDI tool in the international MICS surveys implies that despite obvious cultural and linguistic differences, all children globally should be measured in a similar fashion. Further research is needed on whether the ECDI literacy domain is relevant in this context.

A high proportion of children in our cohort were stunted—50.5% had moderate or severe and 19.0% were severely stunted. Stunting is so closely associated with developmental risk that it is used as a proxy in international estimates(1). In this setting of very high stunting rates, chronic malnutrition was not statistically significantly associated with developmental risk when controlling for other factors. In an analysis of MICS data from multiple countries, stunting was observed to be a good predictor for some, but not all domains of the ECDI; the relationship between stunting and literacy-numeracy varied by country and high vs. low breast feeding status(Miller et al 2015). These results highlight the importance of the PNAN III strategy for reducing malnutrition in these areas, as many cognitive effects of chronic malnutrition may only become apparent as deficits in metacognitive skills (such as memorization, concentration and attention) at a later age when a child starts school and learning may be compromised.

Malnutrition reduction must include not just prevention of new cases but effective treatment of prevalent cases to assist in counteracting growth faltering. In other areas with high rates of stunting, robust malnutrition interventions targeted at smaller areas have been successful at reducing stunting at the population or community level(10, 26). However, important hidden factors contributing to growth-faltering, such as gender roles, age at onset of stunting, breast feeding duration, maternal mental health, dietary diversity and micronutrient availability can vary from site to site; thus formative research in communities should be undertaken to understand how national programs can be adapted on bolstered to have greatest effects.

Adult disengagement was also quite high in this cohort, with almost 25% of children getting no stimulating activities at all from adults. In this setting, engagement by an adult other than the mother or father was quite important. Unfortunately, the standardized questionnaire from the MICS (which we used in this study) does not indicate the specific relationship of the other adult to the child, so we were unable to identify which other adult engaged with the child. Many households in Madagascar as well as in this cohort include multiple generations. The median number of people per household in our cohort was 5, with a range from 1 to 18. Children in Ifanadiana, especially those born to young or unmarried mothers, are frequently raised in a grandparent-headed household. When a mother marries, she may take her young children with her, but often they are left in the household of the maternal grandparents (FR, personal communication), and in some areas of Madagascar, families practice a custom of giving the firstborn child to a grandparent to raise(LRR, personal communication). Grandmothers or aunts, thus, are often responsible for young children, especially those of younger mothers. It is reasonable to suppose that most often, a grandmother is the most deeply engaged “other” adult. Evidence-based interventions that incorporate both nutrition and development stimulation activities could be developed or adapted using community-based participatory research in this setting.

Although only 5% of our population were born to adolescent mothers, being under 20 at the birth of the child was associated with delay compared to both international standards and to peers. Around the world, mothers play a central role in the development of their children. In Madagascar, as in many countries, socialization and discipline of a child until approximately age 5 is generally the purview of the mother or other female relatives (27). Early motherhood is associated in global studies with infant and child mortality, preterm birth and stunting, both of which are associated with reduced child development ((28, 29)). Girl child marriage (defined as marriage before the age of 18) has been associated with off-track development in multiple countries including Madagascar; much of this association is explained by disparities in advanced education and wealth (30). Supporting young women to marry and initiate childbirth later could, thus, have intergenerational benefits. In Madagascar, the legal minimum age of marriage is 18 years, but in rural areas especially, marriage and childbirth often happen earlier for a variety of cultural and economic reasons. This puts both the adolescent girls who become mothers and their babies at increased risk for worse economic, health and development outcomes. Additionally, understanding of both men’s and women’s fertility preferences can be combined with family planning and ready contraception availability in health centers. Such projects are underway in Ifanadiana through the efforts of PIVOT and the MOH.

Factors such as higher number of paternal supportive activities (and not being left in the care of another child were associated with a higher proportion of risk for delay. While this may reflect some sort of detrimental or harsh parenting practices on the part of fathers, a more likely possibility is that if a child is more in need, fathers spend more time focused on the child at risk. Fathers in Ifanadiana are expected to provide for the family and provide some care in times of emergency, but development is not usually their purview (FR, personal communication). However, if a child is faltering in some way, the father may do more activities with the child than a child who is not. The same thing could be true of being left alone with another child. When children are being weaned (age 2, usually because of the advent of another child) even in parent-headed rather than grandparent-headed households, mothers distance themselves and children then become the responsibility of older children in the household (FR, personal communication). This pattern of young children becoming the responsibility of older children around age 2 has been documented in other areas of Madagascar (27) and is common in other settings outside of Madagascar. Children whose development or behavior may be raising red flags in the family may be less likely to be left with other children than those children with development perceived as normal. Surprisingly, having an illness or injury in the last 4 weeks was positively associated with development, which could suggest that active and social children may be more likely to get ill or injured.

This study has several limitations. We conducted a cross-sectional analysis of population-level data to understand the burden of and factors associated with risk of delay in Ifanadiana district. Causal inference cannot be assumed. To the best of our knowledge, no deep ethnographic studies of the population of Ifanadiana exist, so our understanding of the familial dynamics and relationships in Ifanadiana is based on the work of PIVOT’s community health and social work team, which have daily interactions with local families and vulnerable patients. Therefore it is possible that some nuances of relationships have not been well-characterized. The ECDI is a brief population screen meant to provide population level estimates of risk of delay, and not designed to clinically diagnose actual delay or disability in individual children. Also, we rely on self-reported data—the ECDI is administered in an interview to each child’s mother or primary female care-giver, so reports of adult-provided stimulation activities are dependent on the mother’s knowledge, which could lead to misclassification of adult involvement. Nevertheless, the ECDI has been validated on multiple continents and is a standard tool used for a decade in population level surveys to estimate burden of risk of delay, so our findings can be compared to those from Madagascar as a whole and globally. Despite these limitations, our findings contribute to the small body of literature on the burden and risks of developmental delay in low-resource rural settings. Future qualitative explanatory studies including the use of culturally-relevant development screening tools as well as a longitudinal analysis of future waves of the IHOPE cohort data will help to establish directionality of effects and to propose areas of intervention and risk mitigation.

## Conclusions

Although chronic malnutrition was not independently associated with delay risk in this population with high rates of stunting, a high proportion of Malagasy children in this cohort are at risk for developmental delay, specifically in the areas of early literacy and numeracy. A low proportion of children receive developmentally supportive stimulation from adults, but non-parent adults provide more stimulation in general than either mother or father. Stimulation from non-parent adults is associated with lower risk of dela*y*. When compared with their peers, children born to adolescent mothers had a higher risk of delay at 3-4 years of age. Interventions targeting children’s development should be directed at the whole family, including fathers, non-parent adults and older children, as all have important roles in the raising of young children in this setting. Research on the practice of child marriage would be beneficial in this setting as well.

## Data Availability

The datasets used and/or analysed during the current study will be made public at https://dataverse.harvard.edu/dataverse/PIVOT_IHOPE upon publication. Prior to that time, datasets can be requested from the corresponding author.

